# The Effect of SARS-COV-2 Variant on Respiratory Features and Mortality Among Vaccinated and Non-Fully Vaccinated Patients

**DOI:** 10.1101/2022.07.21.22277907

**Authors:** Thomas D. Hughes, Ajan Subramanian, Rana Chakraborty, Shannon A. Cotton, Maria Del Pilar Giraldo Herrera, Yong Huang, Natalie Lambert, Melissa D. Pinto, Amir M. Rahmani, Carmen Josefa Sierra, Charles A. Downs

## Abstract

**Background:** SARS-CoV-2 (COVID-19) has caused over 80 million infections and 973,000 deaths in the United States, and mutations are linked to increased transmissibility. This study aimed to determine the effect of SARS-CoV-2 variants on respiratory features and mortality and to determine the effect of vaccination status.

**Method:** A retrospective review of medical records (n=63,454 unique patients) using The University of California Health COvid Research Data Set (UC CORDS) was performed to identify respiratory features, vaccination status, and mortality. Variants were identified using the CDC data tracker.

**Results:** Increased odds of death were observed among those not fully vaccinated (Delta OR: 1.64, *p* = 0.052; Omicron OR: 1.96, *p* < 0.01). Later variants (i.e., Delta and Omicron) demonstrated a reduction in the frequency of lower respiratory tract features with a concomitant increase in upper respiratory tract features. Vaccination status was associated with survival and a decrease in the frequency of many upper and lower respiratory tract features.

**Discussion:** SARS-CoV-2 variants show a reduction in lower respiratory tract features with an increase in upper respiratory tract features. Being fully vaccinated results in fewer respiratory features and higher odds of survival, supporting vaccination in preventing morbidity and mortality from COVID-19.

## Introduction

The coronavirus disease of 2019 (COVID-19) pandemic caused by the novel SARS-CoV-2 virus was present on the west coast of the United States in late 2019. Since, there have been multiple waves of infection and ongoing viral mutations. According to the United States Centers for Disease Control and Prevention (CDC), as of April 2022, approximately 80 million people have contracted SARS-CoV-2 and 973,000 people have died [1]. The initial virus, termed the Founder variant, has mutated multiple times into other variants namely Alpha, Delta, and Omicron. With each mutation, the virus has become increasingly transmittable, and associated with a reduction in hospitalizations and fewer severe lower respiratory tract issues, the primary cause of COVID-19 deaths, compared to the Delta variant [2].

Despite virus mutation, vaccination remains the best protection against hospitalization, death, and lessens the risk for long-COVID. According to the World Health Organization, approximately 63% of the US population is fully vaccinated. Similarly, 66-67% of the population in the United Kingdom, Canada, and Australia are fully vaccinated [3]. There are several reasons why vaccinate uptake in the US is low. Barriers to vaccination can be structural or attitudinal [4]. Structural barriers relate to access issues (e.g., cost, convenience, and supply chain issues), whereas attitudinal barriers are associated with low perceived risk of acquiring the disease or potential severe consequences from the disease, perceived risks of the vaccine, lack of trust of agencies that are responsible for the development and distribution of vaccines, misinformation, or misconceptions. Therefore, ongoing research is necessary to confront the attitudinal barriers members of the population may have to promote increased vaccine uptake to the target of 80-85% to prevent and mitigate the ongoing significant morbidity and mortality, disruptions to society, and long-term health consequences.

Severe COVID-19 infection may lead to viral pneumonia and acute respiratory distress syndrome (ARDS). One study by Wu et al. (2020) found that 42% of patients with COVID-19 pneumonia developed ARDS [5]. In addition to acute complications such as ARDS, a recent publication from the National Health Service in the United Kingdom (NHS) suggested that chronic cough, respiratory fatigue, and fibrotic lung disease complicate long-term recovery from COVID-19 [6]. The National Health Service (2020) examined long-term symptoms of diseases caused by other coronaviruses, such as severe acute respiratory syndrome (SARS) and Middle East respiratory syndrome (MERS) and found that up to 30% of patients had persistent lung abnormalities after recovering from the acute illness stage [7]. Furthermore, a study by Huang et al. (2021) linked respiratory symptom clusters with a higher risk of long-term COVID-19 or long-haul COVID-19 [8]. Collectively, these studies illustrate the importance of the respiratory system in SARS-CoV-2 infection and subsequent COVID-19 diseases.

The purpose of this study was to assess the prevalence of upper and lower respiratory tract symptoms across SARS-CoV-2 variants, to determine the effect of vaccination status on the symptoms, and to evaluate the risks of mortality based on variant type and vaccination status. A retrospective review of 63,454 medical records from hospitalized patients with confirmed SARS-CoV-2 infection within the University of California Health COvid Research Data set (UC CORDS) was performed.

## Methods

### University of California Health COvid Research Data Set (UC CORDS)

The UC CORDS comprises de-identified health data across all facilities in the University of California (UC) Health system, encompassing 19 health professional schools, five academic medical centers, and 12 hospitals [9]. It contains the records of more than 700,000 patients with de-identified information to enable safe and secure clinical research. This dataset contains data from both hospitalized patients and outpatients. This study was deemed exempted from obtaining ethical approval by the University of California, IRB. All methods were carried out in accordance with relevant university guidelines and regulations. The UC CORDS dataset contains de-identified data from individuals seeking care in the University of California Health System, as such the Institutional Review Board at the University of California-Irvine, waived the need for obtaining informed consent. No experimental protocols were used in this study of de-identified data; therefore, no approvals were sought from the University.

### Variant and Vaccination Status

The UC CORDS data set did not report variant type; therefore, variants were identified based upon dates when each variant was dominant as reported by the CDC data tracker [1]. Accordingly, the date ranges extended from 01/01/2020 to 06/30/2020 for the Founder variant, 06/30/2020 to 05/31/2021 for the Alpha variant, 07/01/2021 to 11/30/2021 for the Delta variant, and beginning 12/01/2021 for the Omicron variant. Patients who received at least two doses of the vaccine before their positive test result were considered fully vaccinated. Patients who did not receive at least two doses of the vaccine before their positive test result were considered not fully vaccinated (i.e., those who were only partially vaccinated (one dose) or did not receive any vaccinations).

### Inclusion and Exclusion Criteria and Sample

The study involved review of EHR data in the UC CORDS data set. Inclusion criteria for this study included all patients, regardless of age, who had a positive test anywhere in the hospital setting (i.e., emergency department, intensive care unit, or any other hospital unit). For any given positive test, respiratory features data was included in a window of five days prior to the test result and up to 30 days after a positive RT-PCR test for SARS-CoV-2. Exclusion criteria for the study included those whose data was obtained from non-hospital (i.e., clinic-based) outpatient settings and those who had a positive RT-PCR SARS-CoV-2 test outside of the predetermined window.

### Respiratory Feature Identification and Extraction

The 40 most reported features across all body systems were extracted from each variant. Each feature was represented by an ICD-10 code and was subsequently translated into medical terminology. Of these most reported features across each variant, the respiratory features were identified and further categorized into lower and upper respiratory features. The reported frequency of each feature was then normalized per 100 cases.

### Statistical Analysis

Statistical analysis was performed using odds ratios to determine the risk of death for each variant while accounting for vaccination status, and the two-tailed Chi-square test to study the effect of respiratory symptoms of COVID-19 on vaccination status across variants. Chi-square tests are used to study the relationship between categorical variables using a contingency matrix. The tests compared the relationship between the frequency of patients who did and did not report a particular symptom. The contingency matrices were created for each variant separately and they each compared the frequency of a particular symptom between fully and not fully vaccinated patients. A *p*-value of < 0.05 was considered statistically significant. Analyses were conducted using Python (version 3.6) and the SciPy package (version 1.8.0).

## Results

### Demographics and Respiratory Feature Frequency

Table 1 provides demographic information for the 63,454 patients included based on variant (Founder (n= 3,465), Alpha (n= 26,274), Delta (n= 7,786), and Omicron (n=27,633)) and vaccine status (fully vaccinated (n= 7,997), not fully vaccinated (n= 55,573)). Figure 1 shows the normalized cases of aggregated upper or lower respiratory tract features based on the variant and vaccination status. Upper respiratory tract features included acute pharyngitis, acute upper respiratory tract infection, cough, disorder of nasal cavity, and nasal congestion. Lower respiratory tract features included abnormal lung findings, acute respiratory failure, dyspnea, hypoxemia, and pneumonia. Figures 2 and 3 show the normalized cases of upper and lower respiratory features based on variant and vaccination status, respectively. Of note, the frequency of lower respiratory tract features decreased with successive variants while upper respiratory tract features increased.

**Table 1.** Demographic Information (n = 65,158 SARS-CoV-2 infections

|  | Fully Vaccinated |  |  |  |  | Not Fully Vaccinated |  |  |  |
| --- | --- | --- | --- | --- | --- | --- | --- | --- | --- |
|  |  | Founder<br>(n= 0) | Alpha<br>(n = 67) | Delta<br>(n = 1200) | Omicron<br>(n = 6774) | Founder<br>(n = 3465) | Alpha<br>(n = 26207) | Delta<br>(n = 6586) | Omicron<br>(n = 20859 ) |
| **Age, in years (mean, standard deviation)** |  | N/A | 61.25 ± 15.8 | 51.17 ± 18.4 | 45.27 ± 18.5 | 48.65 ± 19.9 | 46.72 ± 20.8 | 39.62 ± 21.6 | 38.54 ± 23.4 |
| **Gender (n, %)** | Female | N/A | 38<br>(56.72) | 650<br>(54.17) | 3911<br>(57.74) | 1696<br>(48.95) | 13553<br>(51.72) | 3378<br>(51.29) | 11340<br>(54.37) |
|  | Male | N/A | 29<br>(43.28) | 547<br>(45.58) | 2834<br>(41.84) | 1769<br>(51.05) | 12647<br>(48.26) | 3205<br>(48.66) | 9496<br>(45.52) |
|  | Unknown or Other | N/A | 0 | 3 (0.25) | 29<br>(0.43) | 0 | 7<br>(0.02) | 3 (0.05) | 23 (0.11) |
| **Race (n, %)** | White | N/A | 31<br>(48.44) | 659<br>(58.16) | 2986<br>(46.69) | 958<br>(29.3) | 8201<br>(33.47) | 2548<br>(41.87) | 7102<br>(37.43) |
|  | Hispanic or Latino | N/A | 15<br>(23.44) | 205<br>(18.09) | 1366<br>(21.36) | 1585<br>(48.47) | 10184<br>(41.57) | 1770<br>(29.08) | 5797<br>(30.55) |
|  | Asian | N/A | 9 (14.06) | 118<br>(10.41) | 939<br>(14.68) | 247<br>(7.55) | 2090<br>(8.53) | 418<br>(6.87) | 2168<br>(11.43) |
|  | African-American | N/A | 2 (3.12) | 47 (4.15) | 411 (6.43) | 175<br>(5.35) | 1576<br>(6.43) | 616<br>(10.12) | 1589<br>(8.38) |
|  | Native Hawaiian or Pacific Islander | N/A | 2 (3.12) | 10 (0.88) | 81 (1.27) | 20 (0.61) | 177 (0.72) | 39 (0.64) | 126 (0.66) |
|  | American Indian or Alaska Native | N/A | 0 | 1 (0.09) | 15 (0.23) | 8 (0.24) | 59 (0.24) | 23 (0.38) | 51 (0.27) |
|  | Unknown or Other | N/A | 8 (11.94) | 160<br>(13.33) | 976<br>(14.41) | 472<br>(13.62) | 3920<br>(14.96) | 1172<br>(17.8) | 4026<br>(19.3) |

**Figure 1.**
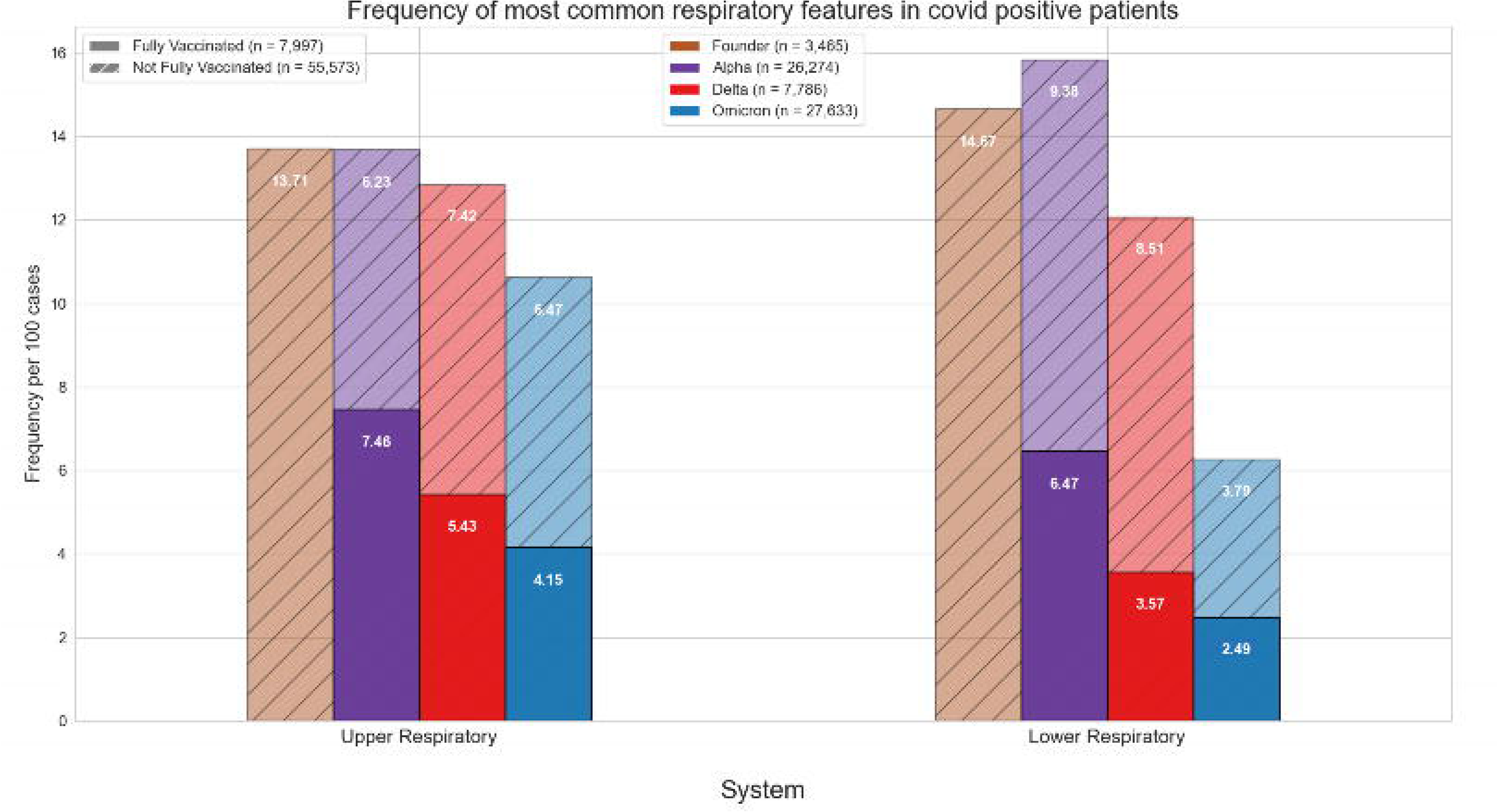
Frequency of Most Common Respiratory Features in COVID Positive Patients. Bar graphs show the frequency of the aggregate upper and lower respiratory features for each variant. Brown color represents the Founder, purple represents the Alpha, Red represents the Delta, and blue represents the Omicron. The bottom bar (solid color, no hatch marks) corresponds to fully vaccinated patients, and the top bar (hatched with diagonal lines) corresponds to the not fully vaccinated patients.

**Figure 2.**
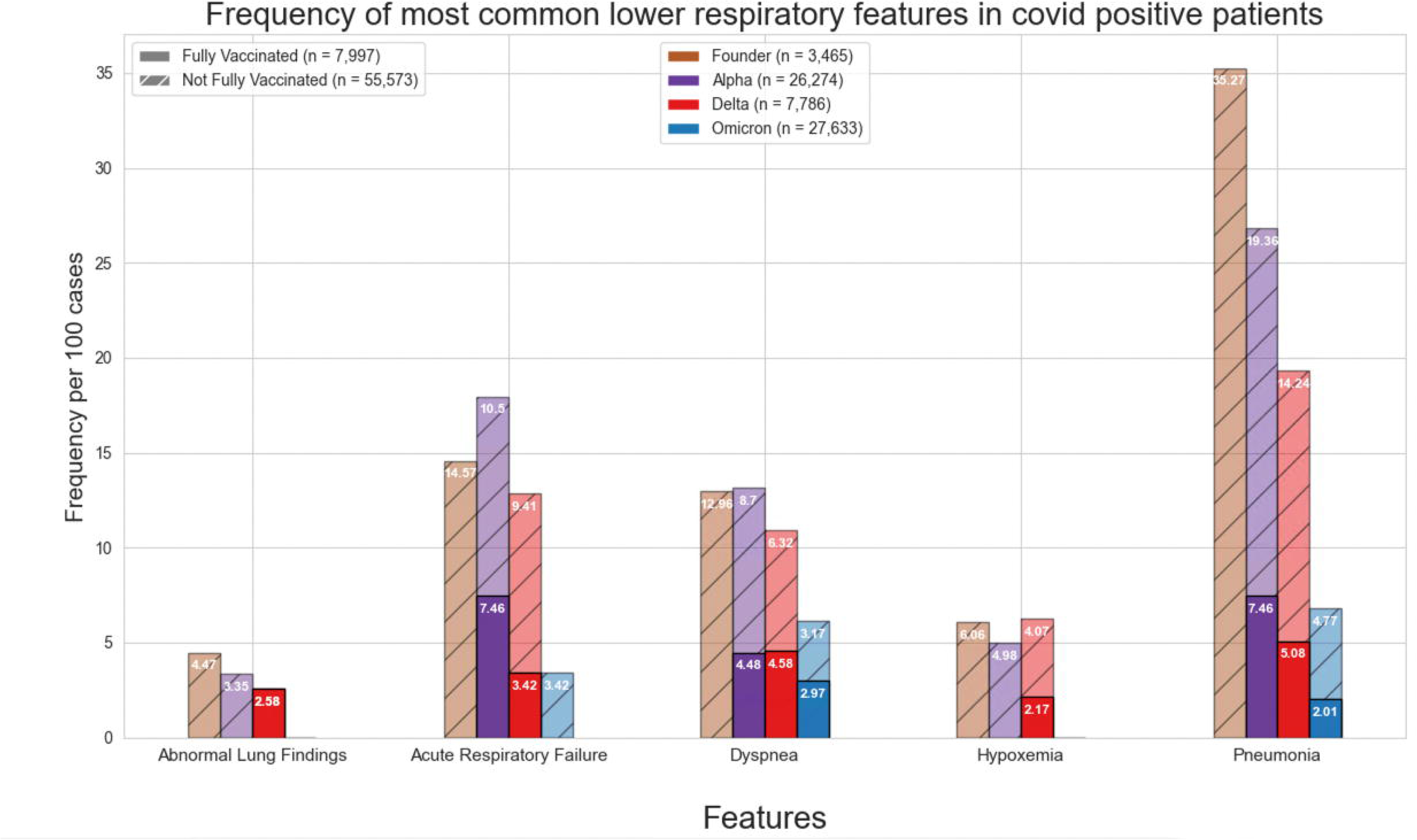
Frequency of Most Common Upper Respiratory Features in COVID Positive Patients. Bar graphs show the frequency of the aggregate upper respiratory features for each variant. Brown color represents the Founder, purple represents the Alpha, Red represents the Delta, and blue represents the Omicron. The bottom bar (solid color, no hatch marks) corresponds to fully vaccinated patients, and the top bar (hatched with diagonal lines) corresponds to the not fully vaccinated patients.

**Figure 3.**
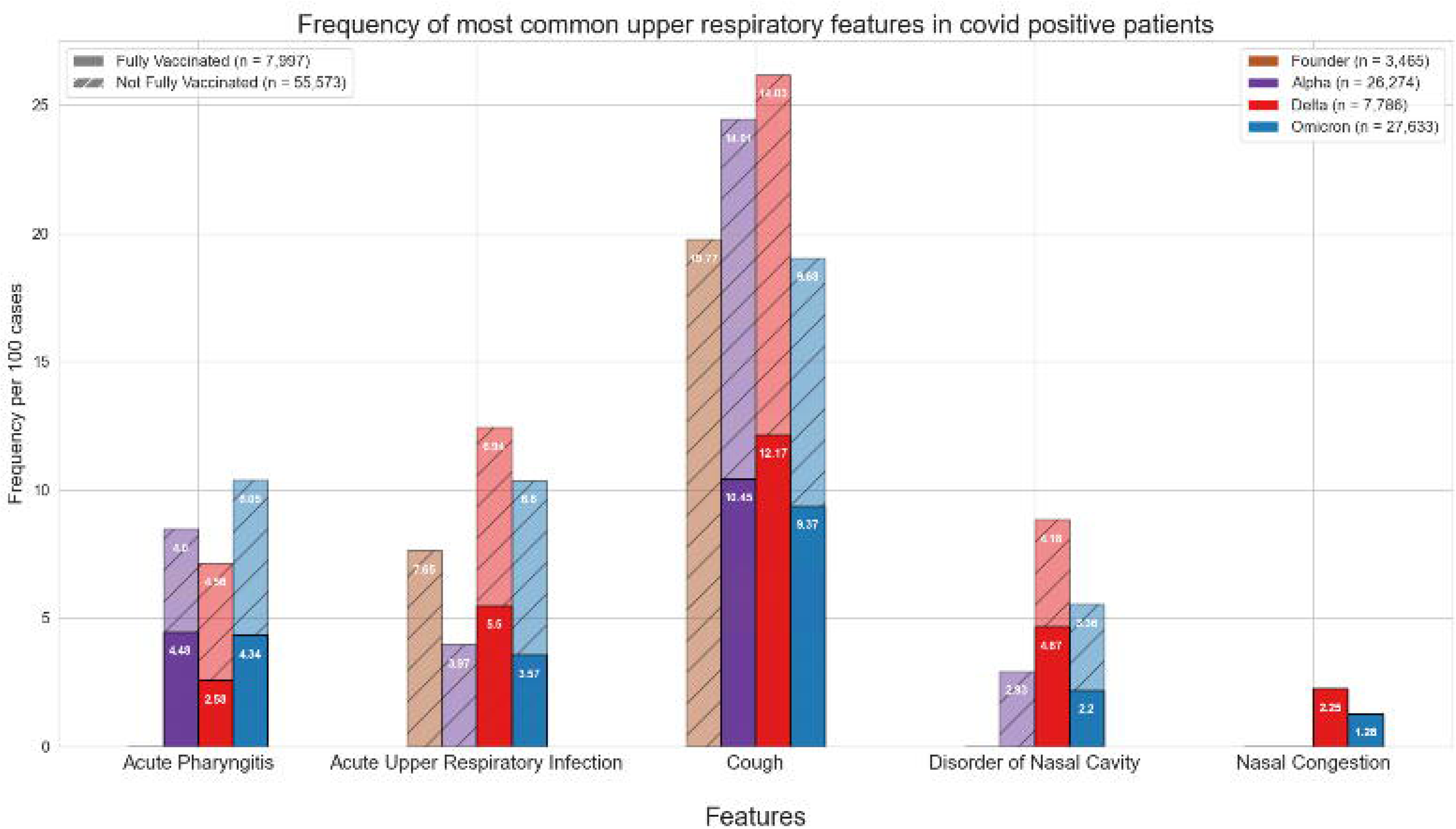
Frequency of Most Common Lower Respiratory Features in COVID Positive Patients. Bar graphs show the frequency of the aggregate lower respiratory features for each variant. Brown color represents the Founder, purple represents the Alpha, Red represents the Delta, and blue represents the Omicron. The bottom bar (solid color, no hatch marks) corresponds to fully vaccinated patients, and the top bar (hatched with diagonal lines) corresponds to the not fully vaccinated patients.

### Mortality

Table 2 provides mortality data for all patients in the study across the four major variants. Patients who were not fully vaccinated had a significantly higher likelihood of mortality during the Delta and Omicron periods compared with those who were fully vaccinated (Delta OR: 1.64, *p* = 0.052; Omicron OR: 1.96, *p* < 0.01). Although the difference in mortality during the Alpha phase did not reach statistical significance, there remained a large difference in the number of patients who died who were fully vaccinated compared with those who were not fully vaccinated (3 versus 997, respectively, *p* = 0.742).

**Table 2.**
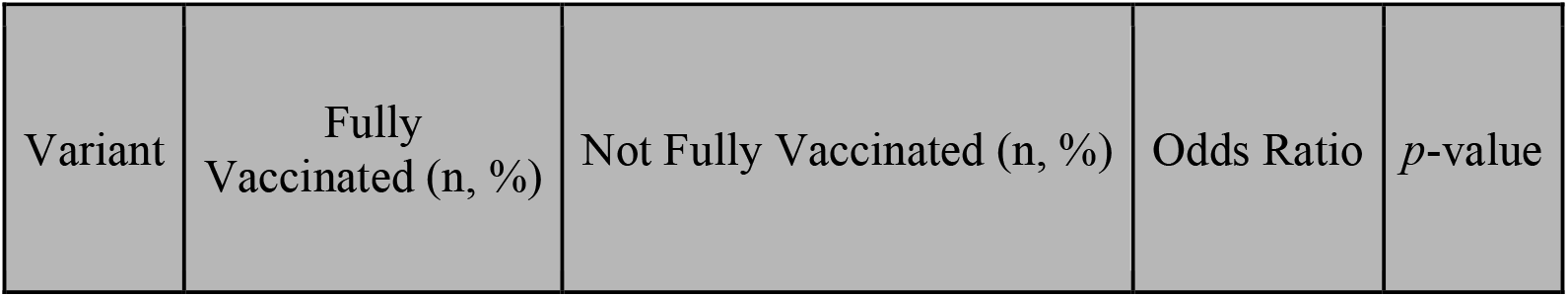
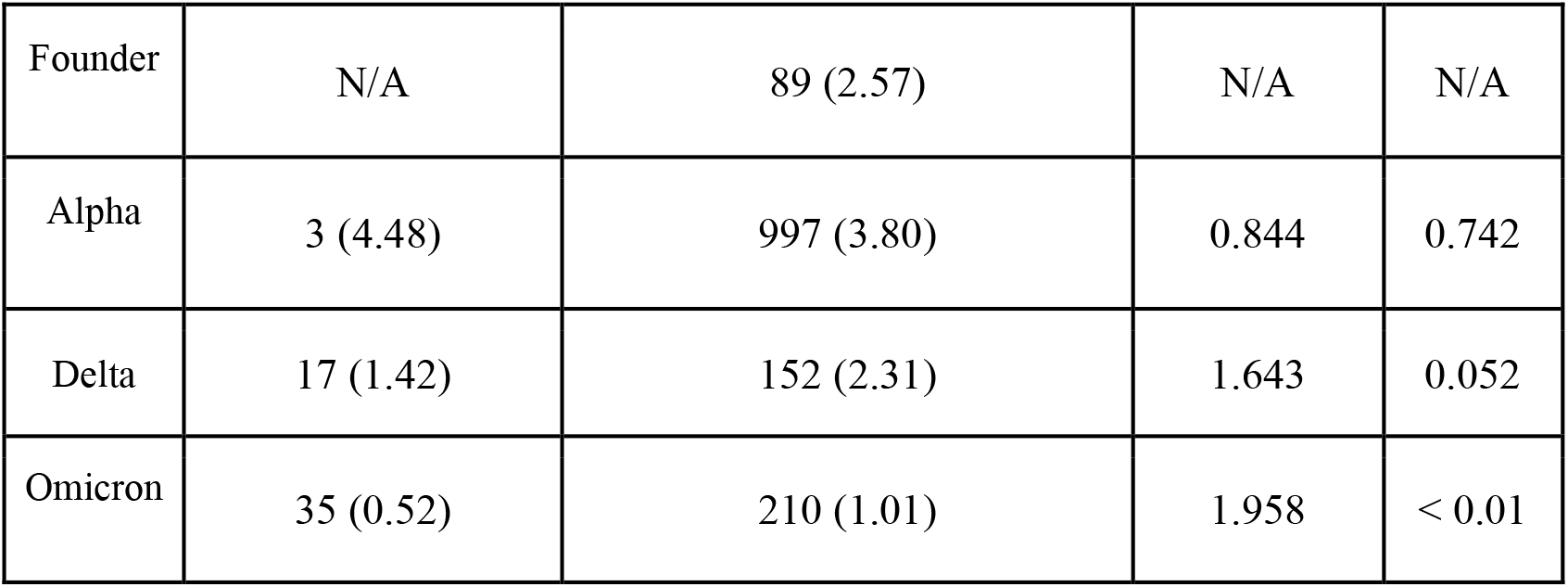
Mortality Rate and Odds Ratio By Variants. p-value < 0.05 is significant.

### Effect of Vaccination Status

The effect of vaccine status on respiratory features was assessed. A total of 7,997 patients obtained two doses of a vaccine and were considered fully vaccinated and 55,573 either received one dose of a vaccine or no vaccine and were not considered fully vaccinated. Of note is that a vaccine was not available until the emergence of the Alpha variant, of which only 67 patients were considered fully vaccinated. Vaccination rates have increased and as a result the ratio between vaccinated and not fully vaccinated individuals is more robust for the Delta and Omicron variants.

We assessed the effect of vaccination status on the frequency of respiratory features based on each variant. Table 3 shows the effect of being fully vaccinated on respiratory features within each variant. These data show that for the Alpha variant being fully vaccinated significantly reduced pneumonia frequency. Similarly, positive vaccination status significantly reduced many lower respiratory tract features including pneumonia, hypoxemia, and acute respiratory failure for those infected with either the Delta or Omicron variants.

**Table 3.** Chi-square Tests Comparing Feature Significance in Vaccinated Versus Unvaccinated Patients. *p*-value < 0.05 is significant

| Variant | Feature | Fully Vaccinated (n) | Not Fully Vaccinated (n) | Chi-square *p*-value |
| --- | --- | --- | --- | --- |
| Alpha | Pneumonia | 6 | 5796 | 0.0144 |
| Delta | Dyspnea | 55 | 416 | 0.0244 |
|  | Acute Respiratory Failure | 50 | 744 | < 0.01 |
|  | Pneumonia | 75 | 1096 | < 0.01 |
|  | Hypoxemia | 26 | 268 | < 0.01 |
|  | Acute Pharyngitis | 31 | 300 | < 0.01 |
| Omicron | Acute Respiratory Failure | 90 | 853 | < 0.01 |
|  | Pneumonia | 168 | 1285 | < 0.01 |
|  | Hypoxemia | 67 | 317 | < 0.01 |
|  | Acute Upper Respiratory Infection | 242 | 1418 | < 0.01 |
|  | Acute Pharyngitis | 294 | 1262 | < 0.01 |
|  | Disorder of Nasal Cavity | 149 | 701 | < 0.01 |

## Discussion

Acute COVID-19 infection causes numerous respiratory disorders, and as such, it is necessary to investigate its impacts across the respiratory system as new variants have emerged. However, because of the predilection for SARS-CoV-2 in causing impairments to the lungs and respiratory system, this study particularly focused on assessing the direct consequences to the respiratory system over the course of the pandemic by examining the frequency of the most common features related to the most dominant and prevalent SARS-CoV-2 variants. Additionally, this study sought to assess the effect of vaccination status on respiratory features. A few observations warrant additional discussion.

First, during the period in which the Delta and Omicron variants were dominant, those patients who were not fully vaccinated had significantly higher odds of mortality compared with those who were fully vaccinated. This finding corresponds to the findings reported in Table 3, in which there were also significant differences between the fully vaccinated patients and not fully vaccinated patients in the features of pneumonia, hypoxemia, and acute respiratory failure. This finding is consistent with Bouzid et al. (2022) who found that Omicron infection was associated with increased survival compared with the Delta variant [10].

Second, a major source of mortality of COVID-19 disease includes acute respiratory distress syndrome (ARDS) and respiratory failure. We show that as variants evolve there is a reduction in lower respiratory tract features, such as pneumonia, hypoxemia, and acute respiratory failure. This finding may be due to increasing rates of vaccination, a reduction in virulence with successive variants, improvement in management of care for patients with acute COVID-19 infection, acquisition of immunity among those reinfected or a combination of any of these factors. The drastic reduction in the lower respiratory symptoms of pneumonia across variants may best demonstrate this phenomenon-during the Founder phase, pneumonia was reported in 35.27 per 100 cases; however, during the Omicron phase, the frequency of pneumonia was reduced to 2.01 in fully vaccinated patients and 4.77 in not fully vaccinated patients. Moreover, the statistically significant difference between the fully vaccinated and not fully vaccinated patients in frequency of pneumonia supports the evidence regarding the immense benefits associated with vaccination in preventing severe disease.

Third, the respiratory features observed with each variant have evolved. In the current study we observed that the incidence of lower respiratory tract features decreased with successive emergence of variants, and that there was a concurrent increase in upper respiratory tract features. For example, Figure 3 shows a general decrease in frequencies for lower respiratory features with each successive variant. Meanwhile, for upper respiratory symptoms including acute pharyngitis and acute upper respiratory infection, those features have increased from the Delta to Omicron period. The findings are consistent with other studies, such as that by Wang et al. (2022), which showed that the Omicron variant was associated with less likelihood of three-day risk of emergency department visit, hospitalization, intensive care unit admission, and mortality, because of Omicron being less virulent in causing lung-related disease [2].

In contrast to the decreasing lower respiratory symptoms observed in the later SARS-CoV-2 variants, there was a notable increase in the trend of upper respiratory symptoms in this study. In particular, the features of acute pharyngitis, acute upper respiratory infection, and cough all either increased or remained elevated during more recent stages of the pandemic. These findings suggest that infection with more recent SARS-CoV-2 variants produces more upper airway features than lower airway features. However, features such as disorder of nasal cavity and nasal congestion were two symptoms that did not change. This finding may be related to the unique symptomatology of the Omicron variant; it may affect the throat and upper respiratory tract more than the nose itself. More research is necessary to conclude that this is the case, but these findings are congruent with studies involving animals [11-12].

### Limitations

This study is not without limitations. First, the nature of the retrospective design is biased towards those who either sought care or required hospitalization for COVID-19; thus, these data will not include information regarding patients who did not seek care. Second, this study did not account for the timing of when patients received vaccination and when they became ill with COVID-19. The possibility remains that patients may have received the full doses of the vaccine, but perhaps developed COVID-19 prior to the time required by their body to develop sufficient antibodies. Third, this study did not account for demographic factors or comorbid conditions of the included patients; it is well-known that those who are older and have pre-existing conditions are more likely to develop worse outcomes than those who are generally younger and healthier. Fourth, although the UC CORDS dataset contains the records of over 2,500 patients infected with the Founder variant, at this stage of the pandemic, the virus was still a novel phenomenon and the UC CORDS database had not yet been fully set up; therefore, the records of some of the patients infected with the Founder variant may be lacking or missing. Fifth, symptom selection was determined through expert identification of symptoms and corresponding ICD-10 code obtained from the electronic health record, so there may be diagnoses which existed but were not captured in the UC CORDS database. Moreover, both “pneumonia” and “viral pneumonia” ranked in the top 40 of feature collection, but due to similarities in presentation, “viral pneumonia” was combined with pneumonia, thus raising the possibility that the actual frequency of pneumonia may have been slightly lower than what was captured in this study due to potential overlap. Sixth, genomic data of the viral strains is unavailable in the UC CORDS dataset, and therefore it was necessary to rely on the information from the CDC’s data tracker to determine time periods in which a particular strain was most likely to be dominant in the US; it is highly probable that the various strains overlapped as newer strains became more dominant. Seventh, there were likely some patients who developed re-infection resulting in their records to be counted twice leading to inconsistent total number of cases included in the study. Lastly, this study did not consider patients who may have had previous COVID-19 infection or having prior COVID-19 vaccination and a booster dose as having prior immunity; therefore, there may be patients in the not fully vaccinated group who retained some level of immunity related to previous COVID-19 infection.

### Future Research

This study primarily focused on the most common respiratory features of patients with COVID-19 while controlling for vaccination status. Future studies should examine the frequency data while controlling for demographic factors, comorbidities, and in relation to the timing of vaccine administration. Furthermore, because vaccination guidelines changed throughout the pandemic, it is necessary to re-examine the data while using the latest vaccine guidelines (e.g. fully vaccinated in this study was considered to be at least two doses; however, patients may have received either an adenovirus vaccine, which initially only required one dose, while other may have received a mRNA vaccine, which initially required two doses). As guidelines continue to shift and the virus continues to mutate, future research should take into consideration the changing guidelines and definition of what “fully vaccinated” is considered to mean.

Additionally, as ongoing research demonstrates the significant long-term effects of COVID-19 infection on developing post-acute sequelae of SARS-CoV-2 infection (PASC), it is imperative to assess how acute COVID-19 infection manifests for patients in the long term. Of particular concern is of how the different variants may be associated with the development of PASC symptoms. Finally, as the COVID-19 pandemic continues to cause immense problems for patients and the healthcare system alike, it is essential to examine historical data to help inform present and future decision-making.

## Conclusion

This retrospective study examined the frequency of respiratory features across four major variants since the outset of the COVID-19 pandemic. Additionally, patients were categorized based on vaccination status and mortality risk was assessed. This study found that there were significant reductions in the risks of mortality for patients who were vaccinated during the Delta and Omicron periods. Additionally, there are substantially fewer lower respiratory features associated with later variants, such as Omicron. Meanwhile, as the frequency of lower respiratory features has decreased, there is a substantial uptick in the frequency of upper respiratory features. This study also showed substantial favorable benefits in patients who are fully vaccinated compared with the unvaccinated or only partially vaccinated, the fully vaccinated population experienced significantly fewer features involving the upper and lower respiratory tract. This study indicates that because of numerous factors, including viral evolution, enhanced immunity, and likely improved treatment modalities, respiratory features involving the lower respiratory tract are reported with less frequency compared with earlier stages of the pandemic.

## Data Availability

The dataset used in this study, University of California Covid Research Data Set (UC CORDS), is made available by the University of California Office of the President and the University of California Biomedical, Research, Acceleration, Integration, and Development. The datasets analyzed during this study are not publicly available. Questions regarding the UC CORDS Data Set can be directed to the project contact, Atul Butte.

https://covidresearch.ucsf.edu/projects/uc-health-covid-research-data-set-uc-cords

## Acknowledgements

The authors would like to thank Joseph Wu from the UC CORDS data team who manages the UC CORDS database.

## Author Contributions

AS and TH conceptualized the study. TH, AS, CD, MP, AR, SC, & CS contributed to the writing of the manuscript. MH, NL, & RC contributed to planning and revisions of the manuscript. AS performed primary data retrieval and analysis and YH helped with data retrieval and analysis.

